# Mandibular dose-volume predicts time-to-osteoradionecrosis in an actuarial normal-tissue complication probability (NTCP) model: External validation of right-censored clinico-dosimetric and competing risk application across international multi-institutional observational cohorts and online graphical user interface clinical support tool assessment

**DOI:** 10.1101/2024.08.20.24312311

**Authors:** MD Anderson Head and Neck Symptom Working Group, Laia Humbert-Vidan, Serageldin Kamel, Andrew Wentzel, Zaphanlene Kaffey, Moamen Abdelaal, Kyle B. Spier, Natalie A. West, G. Elisabeta Marai, Guadalupe Canahuate, Xinhua Zhang, Melissa M. Chen, Kareem A. Wahid, Jillian Rigert, Seyedmohammadhossein Hosseinian, Andrew J. Schaefer, Kristy K. Brock, Mark Chambers, Adegbenga O. Otun, Ruth Aponte-Wesson, Vinod Patel, Andrew Hope, Jack Phan, Adam S. Garden, Steven J. Frank, William H Morrison, Michael T Spiotto, David Rosenthal, Anna Lee, Renjie He, Mohamed A. Naser, Erin Watson, Katherine A. Hutcheson, Abdallah S. R. Mohamed, Vlad C. Sandulache, Lisanne V. van Dijk, Amy C. Moreno, Teresa Guerrero Urbano, Stephen Y. Lai, Clifton D. Fuller

## Abstract

**Background:** Existing studies on osteoradionecrosis of the jaw (ORNJ) have primarily used cross-sectional data, assessing risk factors at a single time point. Determining the time-to-event profile of ORNJ has important implications to monitor oral health in head and neck cancer (HNC) long-term survivors.

**Methods:** Demographic, clinical and dosimetric data were retrospectively obtained for a clinical observational cohort of 1129 patients with HNC treated with radiotherapy (RT) at The University of Texas MD Anderson Cancer Center. ORNJ was diagnosed in 198 patients (18%). A multivariable logistic regression analysis with forward stepwise variable selection identified significant predictors for ORNJ. These predictors were then used to train a Weibull Accelerated Failure Time (AFT) model, which was externally validated using an independent cohort of 265 patients (92 ORNJ cases and 173 controls) treated at Guy’s and St. Thomas’ Hospitals.

**Findings:** Our model identified that each unit increase in D25% is significantly associated with a 12% shorter time to ORNJ (Adjusted Time Ratio [ATR] 0·88, p<0·005); pre-RT dental extractions was associated to a 27% faster (ATR 0·73, p=0·13) onset of ORNJ; male patients experienced a 38% shorter time to ORNJ (ATR 0·62, p = 0·11). The model demonstrated strong internal calibration (integrated Brier score of 0·133, D-calibration p-value 0.998) and optimal discrimination at 72 months (Harrell’s C-index of 0·72). The model also showed good generalization to the independent cohort, despite a slight drop in performance.

**Interpretation:** This study is the first to demonstrate a direct relationship between radiation dose and the time to ORNJ onset, providing a novel characterization of the impact of delivered dose not only on the probability of a late effect (ORNJ), but the conditional risk during survivorship.

**Funding:** This work was supported by various funding sources including NIH, NIDCR, NCI, NAPT, NASA, BCM, Affirmed Pharma, CRUK, KWF Dutch Cancer Society, NWO ZonMw, and the Apache Corporation.

## Introduction

Osteoradionecrosis of the jaw (ORNJ) is a severe iatrogenic sequela of radiotherapy (RT) that impacts patients treated for head and neck cancer (HNC) at an estimated prevalence of 4-15% ^1^. Radiation-associated devascularization of bone and normal tissue injury lead to loss of cortical bone integrity which fails to heal, resulting in a constellation of symptoms that substantially reduce quality of life and limit oral function ^2,3^. Depending on severity, ORNJ management ranges from conservative therapy (i.e., topical antimicrobials) to morbid surgical extirpation of nonviable tissue. However, these treatments often target the alleviation of disease symptoms rather than the root causes of ORNJ ^4^. Additionally, the lack of early detection methods and risk assessment tools for ORNJ make prevention and detection difficult for medical professionals to preemptively manage the condition, often delaying care and resulting in costly, invasive interventions.

Patients are at a lifetime risk of developing ORNJ following RT, which is of increased concern for patients who are now living much longer following RT for HNC. The explosive growth of human papilloma virus-associated (HPV+) oropharyngeal cancer (OPC) often affecting non-smokers has resulted in more longterm survivors of patients treated with radiation for HNC. For this reason, determining the long-term (>2 year) time-to-event profile of ORNJ has important implications for professionals who may be monitoring the oral health of long-term survivors for many years after intensive disease surveillance has transitioned to survivorship ^5^.

Numerous studies ^6–9^ have examined the statistical correlation between ORNJ and various dosimetric, clinical, and demographic risk factors. In a previous investigation ^10^, we pioneered and externally validated the first ORNJ Normal Tissue Complication Probability (NTCP) model. While these investigations offer invaluable insights that steer clinical decision-making and treatment plan optimization, they largely rely on cross-sectional datasets, where the potential for reporting bias is extant, and do not account for right-censoring (i.e., consideration of cases without event or who are under surveillance and have not had ORNJ but remain at risk). As Van den Bosch et al. ^11^ note “NTCP-models are generally developed for a single complication grade at a single time point”, thus overlooking the temporal variability in toxicity risk.

Understanding the influence of treatment decisions and risk factors on the timing of ORNJ is crucial for effective prevention and management. Treister et al. ^12^ carried out risk factor association analysis on a longitudinal ORNJ data set with data points at 6, 12 and 24 months and identified pre-RT extractions, higher RT dose and tobacco use as significant risk factors. The present study aims to model longitudinal associations to provide patient-specific ORNJ risk predictions over time. Non-parametric and semi-parametric models, such as Cox proportional hazard, have been widely used. However, these methods may not fully capture the nuanced temporal dynamic of the event due to their broader assumptions about data distribution. Accelerated Failure Time (AFT) models offer a valuable parametric alternative, which enhances the interpretability of each factor’s influence on the event onset. Weibull models are especially attractive as a parametric approach for time-to-event applications for risk-prediction ^13–15^ as an interpretable alternative to the clinically familiar non-parametric proportional hazards methods.

As part of a larger effort to leverage right-censored models to inform risk-based surveillance and prophylactic management trial enrollment ^16^, as well as considering challenges to traditional NTCP models ^17^, we have sought to undertake the following specific aims:

i) Determine relative actuarial incidence of ORNJ over time and establish the relationship between mandibular dose-volume and actuarial progression-to-ORNJ. ii) Derive actuarial NTCP models for ORNJ that incorporate right-censored data for patient risk stratification. iii) Derive and externally validate a dose-aware time-to-event model with clinico-demographic factors. iv) Develop and test an online clinical decision support tool with a graphical user interface (GUI) for clinical implementation of the risk model, including formal stakeholder usability testing.

## Methods

A multivariable time-to-event prediction model was developed on an internal dataset from MD Anderson Cancer Centre (MD Anderson) and externally validated on an independent cohort of a British population treated at Guy’s and St Thomas’ NHS Foundation Trust (GSTT).

### Patient Selection

After institutional review board approval (RCR030800), data from a philanthropically funded observational cohort (Stiefel Oropharynx Cancer Cohort, PA14-0947) were extracted for retrospective acquisition. Patients in the internal MD Anderson cohort included all consented RT cases treated with curative intent from 2005 to 2022. Patients undergoing RT for HNC are closely followed up with clinical and radiological assessments every 3 to 6, 12, 18 to 24 months then approximately annually after the end of the RT course. An external cohort was obtained retrospectively from the HNC clinical database maintained at GSTT under the Northwest - Haydock Research Ethics Committee of the NHS Health Research Authority (REC reference 18/NW/0297, IRAS project ID: 231443); patients treated between 2011 and 2022 were included. The GSTT clinical protocol for HNC patients includes clinical follow up for five years. Control subjects in the GSTT cohort were retrospectively matched with a 2:1 ratio based on primary tumor site and treatment year. Incomplete or not available datasets were excluded.

### Clinical Endpoint

The primary analysis framework focused on ORNJ as the sole event of interest. The primary endpoint was defined as the development of any physician-reported grade of ORNJ following the initiation of RT (i.e., ORNJ vs. no ORNJ), with the time to event (TTE) recorded as the interval (in months) from the start date of RT to the first documented instance of ORNJ in the patient’s electronic medical health record. As the current datasets pre-date recent consensus recommendations ^18,19^ and used then-institutional standard clinical reporting, we designated all cases that were clinically and/or radiographically deemed ORNJ in the MD Anderson and GSTT cohorts as ORNJ and deferred to listed clinical documentation; this was necessary as divergent ORNJ grading systems (Tsai/Notani) were in-use (Appendix B). Patients without confirmed ORNJ diagnosis were right censored at the date of last contact.

### Data & Statistical Analysis

#### Multivariable Logistic Regression Analysis

Clinical and dosimetric parameter were collected from annotated sources at MD Anderson and GSTT using a described methodology and process (Appendix B). Multivariable logistic regression (LR) analysis with forward stepwise variable selection was carried out in R statistical software using the clinical and dosimetric data to identify the most significant variables for prediction of ORNJ to be considered for the Weibull AFT model. The LR analysis and subsequent NTCP modelling was carried as per the methodology previously used by van Dijk et al. (Appendix D).

#### AFT Model Development

A Weibull distribution is characterized by two main parameters: a scale parameter (λ), determining the distribution spread over time, and shape parameter (*ρ*) which indicates whether the rate of the event increases (*ρ* >1), decreases (*ρ* <1), or remains constant (*ρ* =1) over time. Considering covariates X_1_, X_2_, …X_n_, the function of the scale parameter can be expressed as λ(x) = exp(β_0_ + β_l_X_l_ + β_2_X_2_ +··· + β_n_X_n_), where β_0_ is the intercept of the transformed scale when all covariates are at their reference level, while β_1,_ β_2,_ …., β_n_ are the coefficients of the log-linear relationship between each covariate and the time to event ^20–23^. The corresponding survival function for the Weibull AFT model is articulated as 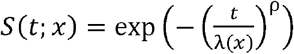 where *S*(*t;x*) represents the probability of a patient surviving beyond time *t* without experiencing ORNJ, given their specific covariates *X*.

Considering β_D25_, β_gender_, and β_dental_ the coefficients of the covariates D25% (*X*_*D25*_), gender (*X*_*gender*_), and pre-RT tooth extraction (*X*_*dental*_), respectively, on the log-transformed time to ORNJ, the function of the scale parameter, pertinent to our study, is thus expressed as λ(x) = exp(β_0_ + β_*XD25*_ *X* _*D25*_ + β_*gender*_*X*_*gender*_ + β_*dental*_*X*_*dental*_).

Adjusted Time Ratios (ATRs), calculated as the exponential of regression coefficients, were used to interpret the proportional impact of the model’s covariates on the time to ORNJ for one unit increase in continuous variables or in relation to reference group(s) in categorical ones.

The analysis and WAFT model development was conducted in Python programming environment (version 3.11) using the ‘*WeibullAFTFitter*’ function of the *Lifelines* (version 0.28) survival analysis library ^24^.

#### AFT Model evaluation

The Weibull AFT model was internally and externally validated. For internal validation, the dataset was randomly split into training (80%) and test (20%) subsets with balanced ORNJ status representation. The Weibull AFT model was fitted to the training data subset and internally evaluated on the test subset. Model performance was assessed in terms of overall performance, predictive accuracy and model calibration on both the internal and external datasets using time-independent metrics ^25^. The Brier score evaluates the mean squared difference between the observed outcomes and the predicted probabilities of event occurrence. In the context of time to event analysis, the Integrated Brier score (IBS) provides a single summary measure of the model’s prediction accuracy over time. The concordance index (Harrell’s C-index) was used to measure the predictive accuracy of the model in terms of its ability to correctly rank the event times. Model calibration was assessed with the Distributional calibration (D-calibration), which is a measure of the calibration of the predicted survival curves (rather than a time-specific outcome prediction).

#### GUI Development and Prospective Assessment

The WAFT-based time-to-ORNJ online calculator graphical user interface (GUI) is available at https://uic-evl.github.io/OsteoradionecrosisVis/ (Figure 1), where the user can either obtain a predicted risk of developing ORNJ at a specific time point or visually assess the time-dependency of ORNJ risk with the different covariates of the ORNJ WAFT model (D25%, gender and pre-RT dental extractions). The usability of the GUI was prospectively evaluated on a test dataset by 25 users of different degrees of expertise and clinical specialties. A Qualtrics survey was designed with eight case-specific questions, the ten questions from the Brooke et al^26^. SUS scale questionnaire and three additional open questions for additional feedback. More details on the survey and the SUS scale are provided in Appendix G.

**Figure 1.**
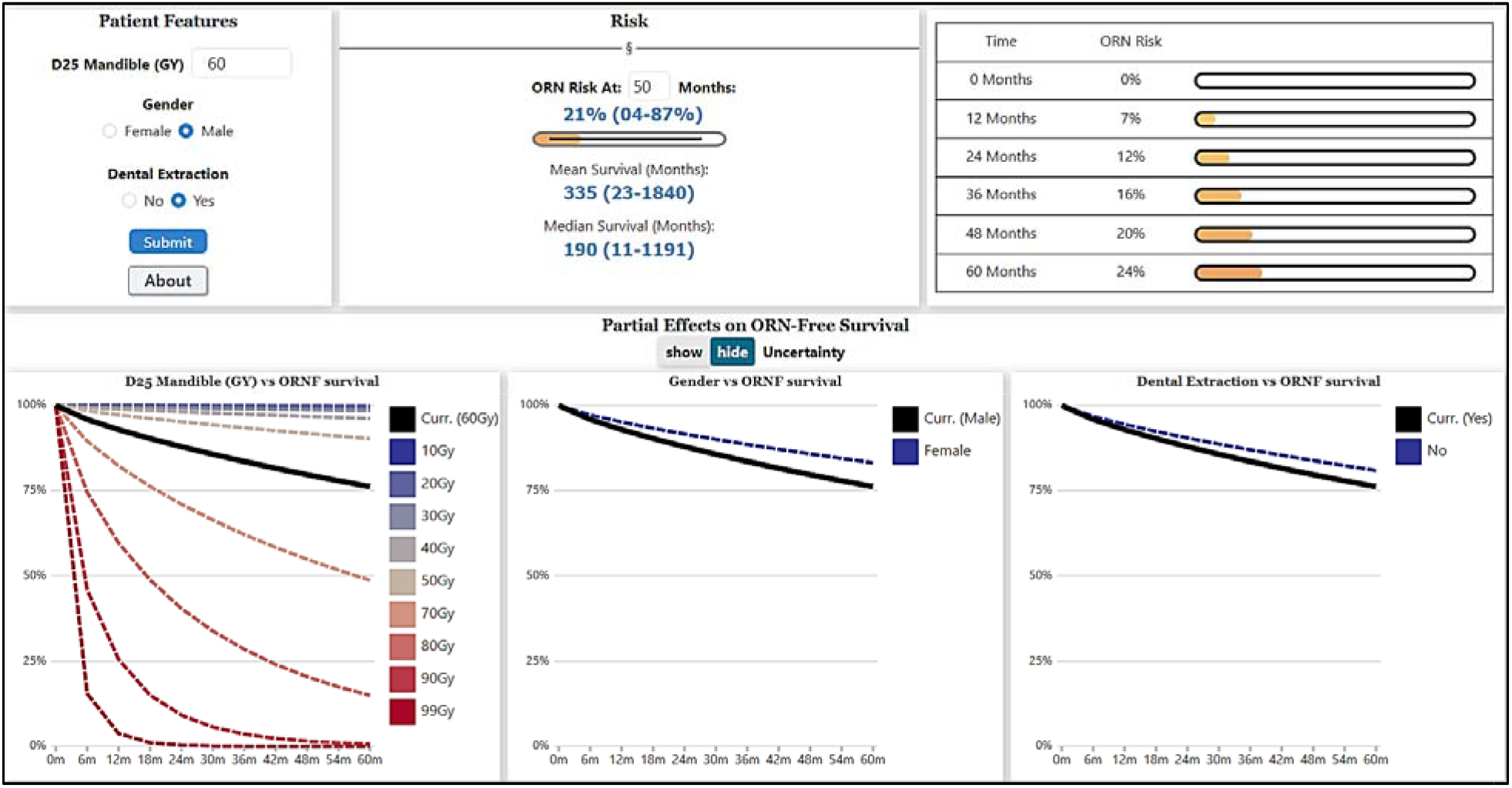
Screenshot of the WAFT-based time-to-ORNJ online calculator GUI.

## Results

### Patient selection

From a population of 1259 MD Anderson patients with HNC, a total of 1129 patients were included in the final analysis, of with ORNJ was observed in 198 cases at the end of follow-up period, with a median time to event of 20·5 months (IQR 35·1). The median follow-up time for the censored group was 71·7 months (IQR 62·7). Actuarial time-to-event (denoting diagnosis of ORNJ as an event censoring at either death or last follow-up) is shown in Figure 2. The external validation GSTT cohort consisted of 92 ORNJ subjects and 173 matched controls. The median time to ORNJ was 13·6 months (IQR 20·3) and the median follow-up time for the control group was 47·3 months (IQR 24·2).

**Figure 2.**
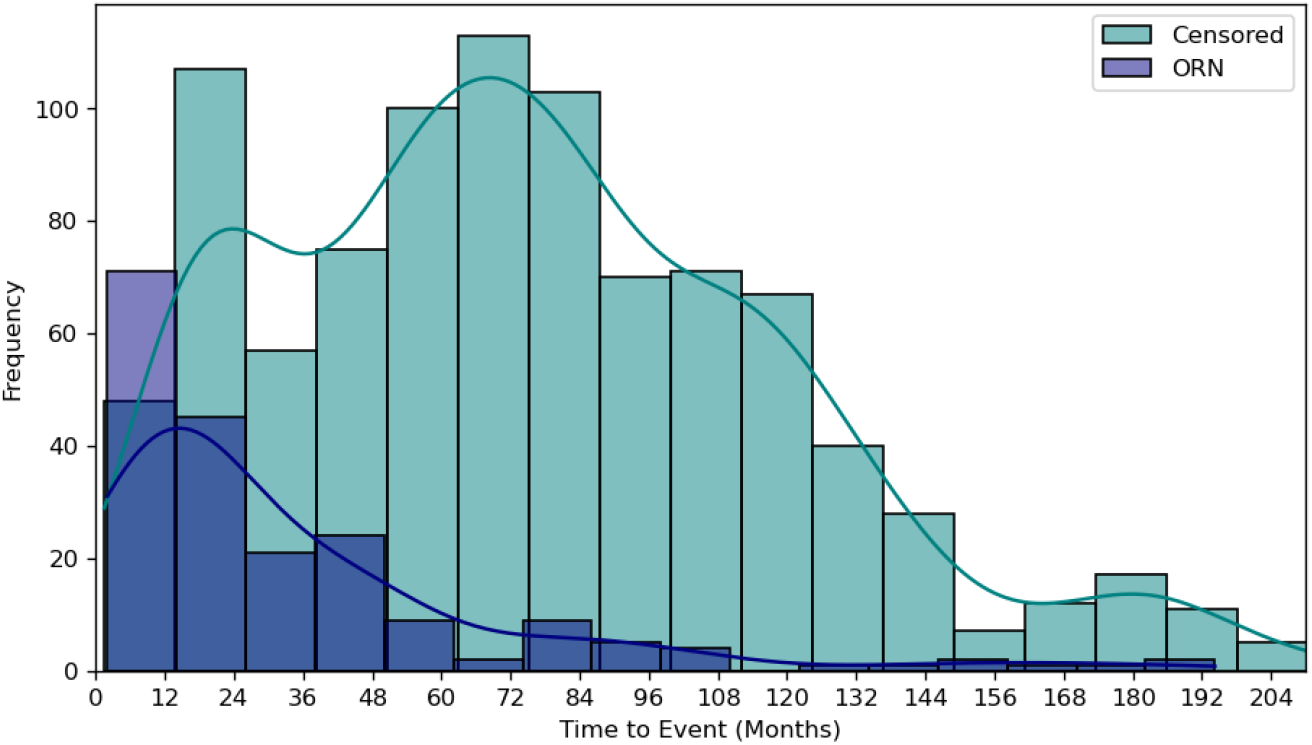
Frequency plot of actuarial time-to-event in months by ORNJ status for the MD Anderson dataset, where diagnosis of ORNJ is considered an event censoring either at death or last follow-up. Appendix C includes frequency plots for the training (Figure C1a), test (Figure C1b) and external (Figure C1c) datasets separately.

### Multivariable Forward Stepwise Logistic Regression Analysis

All DVH metrics and the following clinical variables were considered in the multivariable analysis: smoking status (binary, current vs. never/former), chemotherapy (yes vs. no), gender (binary, male vs. female), pre-RT dental extractions (binary, yes vs. no), post-operative RT (binary, yes vs. no), tobacco pack years (continuous), age (continuous) and mandible volume (continuous).

The forward stepwise multivariable LR process was first applied considering the pre-selected variables. This identified the following dosimetric and clinical variables as the highest predictors of ORNJ: V55Gy(percentage volume of the mandible that receives at least 55 Gy), D25% (minimum dose received by the most irradiated 25% of the mandible volume), pre-RT dental extractions and gender (supplementary Table D1). However, in this first forward stepwise LR process, the model’s coefficient for the V55Gy parameter was largely altered when the D25% parameter was introduced, showing a correlation between the two dosimetric parameters: V55Gy and D25%. Also, the p-value for the V55Gy showed that this parameter was not statistically significant in the prediction of ORNJ. Consequently, the V55Gy parameter was excluded from the pre-selected variables subset and the modelling process repeated on the updated dataset (supplementary Table D2). Therefore, the final ORNJ NTCP model included D25%, pre-RT dental extractions and gender. Details of the ORNJ NTCP model performance are provided in supplementary Table D3.

### Weibull AFT model

The ORNJ Weibull AFT (WAFT) model was trained and tested considering the entire time-to-event range in the MDACC cohort. Details of the WAFT model are provided in Table 1 and the resulting survival curves by variable are represented in Figure 3. The shape parameter of the model (⍰≈ 0·81) indicates a decreasing hazard rate for ORNJ over time among the study group. For the covariates, our findings suggest that each unit increase in D25% is significantly associated with an 12% shorter time to ORNJ (ATR 0·88, p<0·005). We also observed that patients who undergo dental extractions experience ORNJ at a rate 27% faster (shorter time to ORNJ) compared to those who do not undergo pre-RT dental extractions (ATR 0·73, p=0·13). A 38% (ATR 0·62, p=0·11) shorter time to ORNJ was observed in male patients. However, statistical significance of the dental extractions and gender variables was not conclusive. Cumulative hazard function and partial effects of the D25%, pre-RT dental extraction, and gender on survival outcomes were visually analyzed to aid interpretation of the influence of individual factors within the context of the WAFT model (supplementary Figure E1).

**Table 1.** ORNJ WAFT model parameters and coefficients.

| Parameters | Covariates | Coefficients $\beta_i$ (95% CI) | ATR (95% CI) | p-value |
| --- | --- | --- | --- | --- |
| Scale (l) | D25% | -0.12 (-0.15, -0.10) | 0.88 (0.86, 0.91) | <0.005 |
|  | Dental extractions | -0.31 (-0.71, 0.10) | 0.73 (0.49, 1.10) | 0.13 |
|  | Gender | -0.48 (-1.08, 0.11) | 0.62 (0.34, 1.12) | 0.11 |
|  | Intercept | 13.69 (11.77, 15.62) |  |  |
| Shape (r) | Intercept* | -0.21 (-0.35, -0.08) |  |  |

\* The shape parameter ( $\hat{\alpha}$ ) is calculated as the exponential of this intercept, resulting in $\hat{\alpha} = e^{-0.21} \approx 0.81$

**Figure 3.**
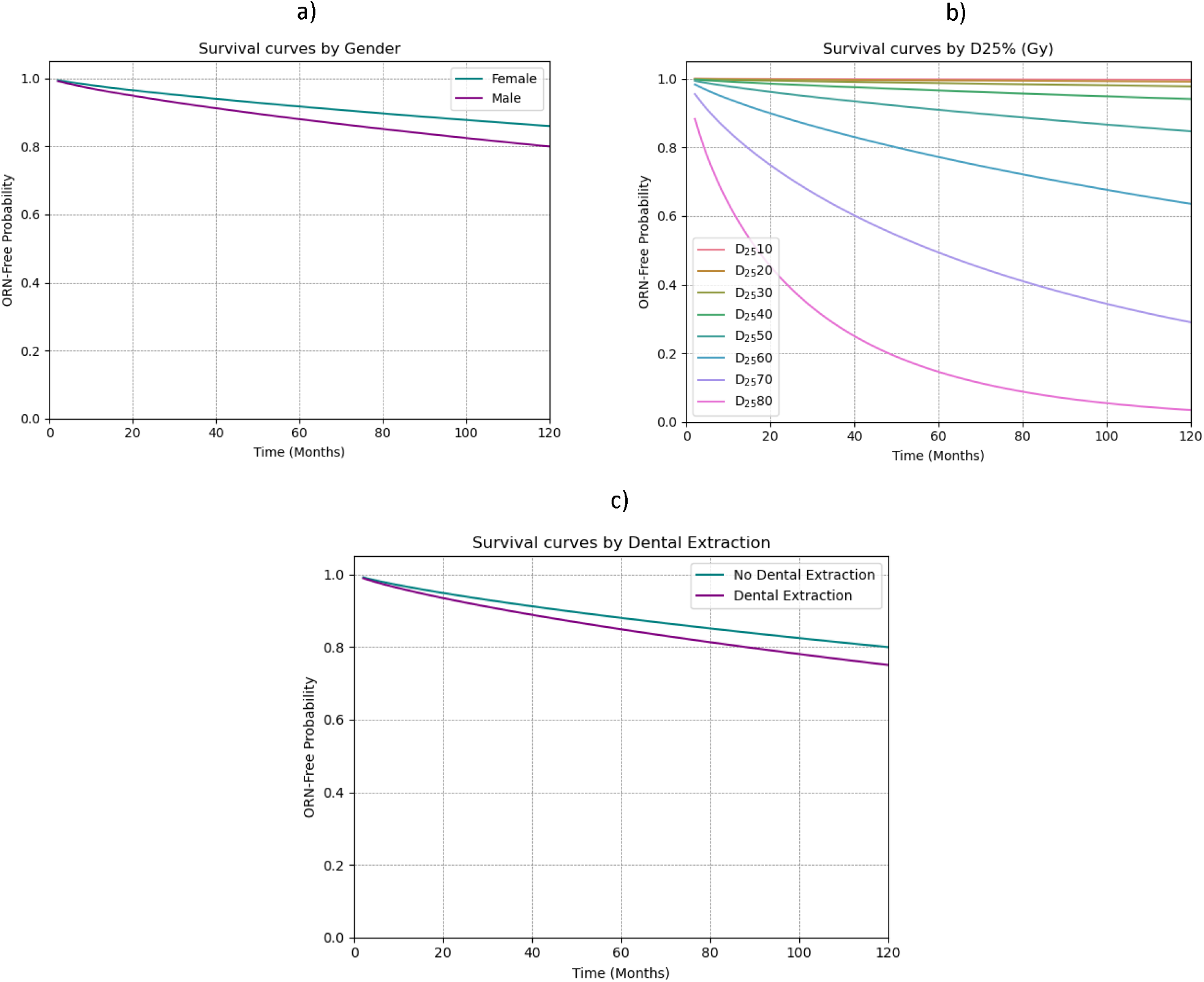
ORNJ WAFT survival curves for the different covariates considered in the model: gender (a), D25% (b) and dental extractions (c).

### Weibull AFT model performance

Maximum discrimination performance of the model at internal validation (Harrell’s C-score of 0·723) was observed at the 72 months predictive horizon (Figure 4a), which coincides with the timepoint where both groups, ORNJ and censored, exhibit the largest difference in the test dataset (supplementary Figure D1b). Model calibration at internal validation was good to excellent, with an integrated Brier score of 0·133 (Figure 4b) and, as shown in Figure 4c, successfully d-calibrated (p-value 0·998 > 0·05). Model performance decreased slightly when tested externally on the independent dataset (Appendix F). The distributional calibration plot (supplementary Figure F1c) shows that the model’s predicted probabilities were consistently low compared to the actual outcomes, i.e., the model was underconfident in its predictions.

**Figure 4.**
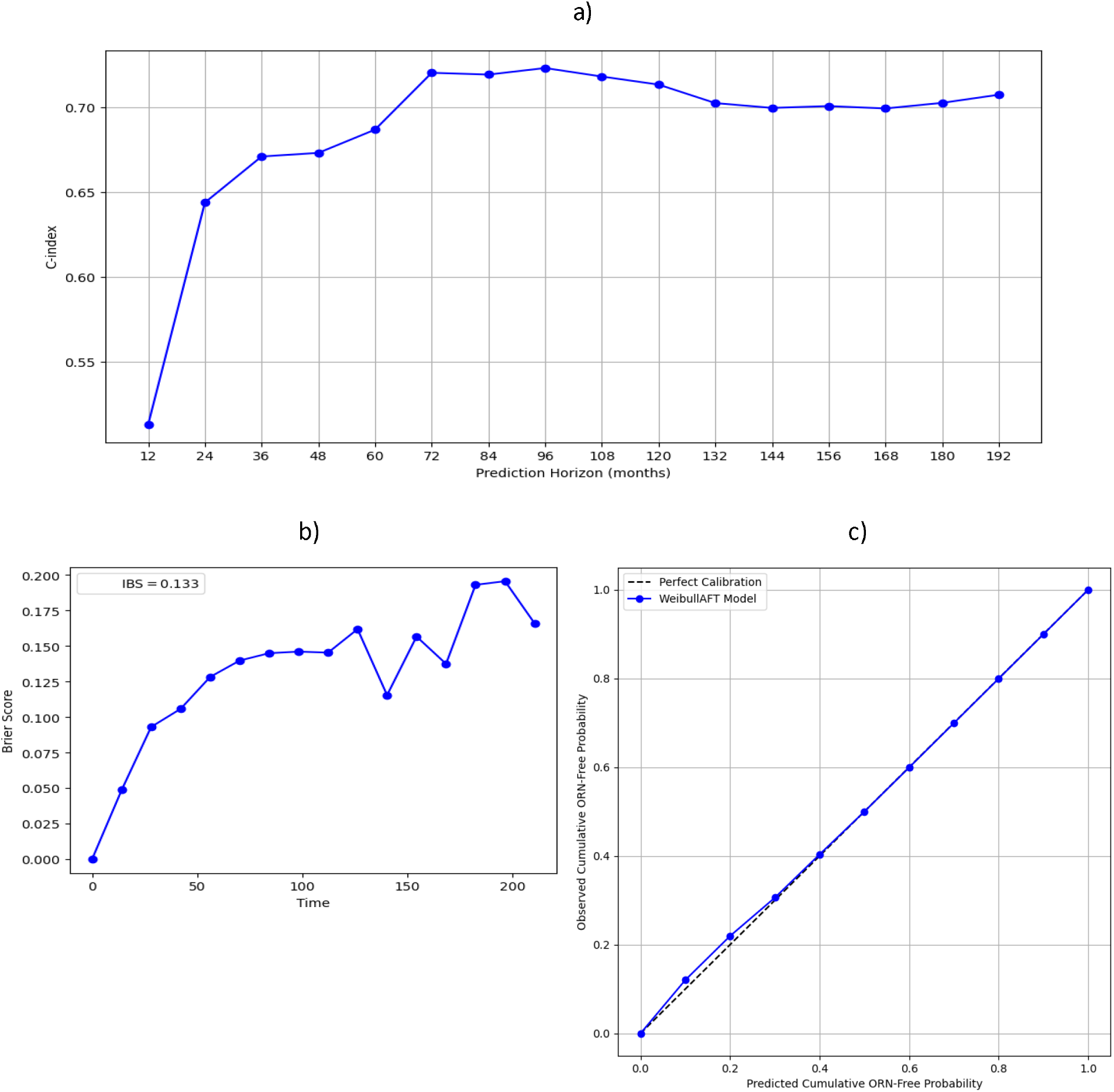
ORNJ WAFT model performance plots at internal validation. Discrimination performance variation over time is described by the Harrell’s C-index (a). Overall model performance over time is described by the Brier score and integrated Brier score (IBS) (b). Model calibration is described by the Distributional calibration curve (c), which describes the computed squared difference between the observed and predicted number of events within different time intervals.

## Discussion

Our work demonstrates for the first time a direct relationship between radiation dose and the time to development of osteoradionecrosis of the jaw (ORNJ) and is a novel characterization of the impact of delivered dose not only on the probability of a late effect (ORNJ), but the conditional risk during survivorship.

Osteoradionecrosis is an orphan disease ^27^ with a currently undefined prevalence, owing to variability in disease classification, and until 2023, lack of a formal International Classification of Disease specific designation [ICD-10 FB81.5], previously denoted as “Other osteonecrosis” without attribution to radiation therapy [ICD-10-CM Diagnosis Code M87.8]. Historically, ORNJ (i.e., FB81.5 Osteonecrosis due to ionizing radiation & Specific anatomy:XA51B7 Mandible) has had over 20 distinct descriptive categories, grading systems, or diagnostic criteria. This ambiguity has led to highly variable estimates of the prevalence of ORNJ, with reports designating between 4-15% ^1^ of HNC RT cases.

The actual time course of development of ORNJ also remains under-described. Using a more restricted criteria of “exposed bone”, the most reliable cross-sectional cohort analysis of post-radiation events ^12^ showed, in 572 longitudinally followed participants, a cumulative rate of exposed bone of 6·1%, with all patients presenting with disease in <18 months; however, this high-quality dataset followed patients only until 24 months post-therapy. Other studies report varying median time-to-event, with clinical features such as oropharyngeal disease site ^28^ or dental extractions ^29^ associated with faster progression to ORNJ. However, formal toxicity modeling of the conditional probability of ORNJ has not been established until now.

In this study, we have successfully developed a novel time-to-event approach for predicting orodental toxicity, thus providing a more comprehensive estimation of disease trajectory to allow effective risk stratification and surveillance strategies. Additionally, we have developed and tested a WAFT-based time-to-ORNJ online calculator graphical user interface (GUI) with overall high usability scoring (Appendix G) that will facilitate clinical implementation of our model.

In a previous study ^10^, a single dose-volume histogram (DVH) parameter-based traditional NTCP model was developed, demonstrating a corollary model using the received dose to 30% or more of mandibular regions of interest (mandible D30%) and pre-radiotherapy dental extraction as predictors of ORNJ. For the current study, prior to the time-to-event model development, we repeated the NTCP modelling exercise after careful manual revision of the dataset. Reassuringly, D25% (close to D30%) and pre-RT dental extractions were also identified as predictors in the updated NTCP model.

Traditional NTCP models not only rely on binarization of the clinical endpoint (e.g., the presence or absence of ORNJ) but also a fixed-interval truncation of surveillance interval without right-censoring. Thus, while there is abundant suggestion that increased radiation dose increases the risk of ORNJ, there is scant data regarding the relative relationship between pre-therapy dose or clinical factors on time to ORNJ development. Consequently, in this study, we aimed to expand on an existing ORNJ NTCP model ^10^ to incorporate the temporal information of ORNJ development, by using a novel application of a prediction model incorporating parametric modeling of continuous right-censored time-to-toxicity-event prediction of ORNJ.

We have previously shown that prediction models that account for right-censoring can provide differential variable selection compared to categorical classification methods ^30^. Reassuringly, the use of right-censor ware TTE models nonetheless also validated our non-right-censored traditional NTCP model, and our results underscore the effect of the risk factors considered (i.e., D25%, dental extractions and gender). Put simply, these factors not only are correlates of ORNJ but are associated with faster interval of ORNJ development. This has important implication for post-RT surveillance in addition to pre-therapy dose reduction strategies. For example, enhanced surveillance imaging methods to monitor progression towards ORNJ, or risk-stratified prevention interventions are potentiated by the proposed model.

By internally and externally validating our model, we have demonstrated its reliability and applicability across diverse patient populations. Despite the slight drop in performance, most likely introduced by differences in treatment protocols and population characteristics, the external validation results indicate that the WAFT model generalized well to unseen data from an independent cohort. As opposed to the internal dataset, the external dataset was a matched cohort, with 2:1 control to case matching based on primary tumor site and treatment year, which could have resulted in a reduced variability of clinical characteristics. Moreover, time to event distributions were very different between the training and the external datasets (supplementary Figure D1): while the model was trained across predictive horizons beyond 204 months, the external test dataset was limited to a maximum of under 100 months. Additional external validation of the WAFT model on a larger and more diverse observational cohort will allow confirmation of the model’s generalizability.

While there are a number of survival functions to be considered for time to ORNJ modelling, our choice of the WAFT model was based on its flexibility and suitability to the study’s objectives. In future work, we will continue to explore comparisons with alternative distribution functions such as the log-linear or exponential distributions.

The presented results focused on a binary endpoint for ORNJ (i.e., any grade of ORNJ vs. no ORNJ). While this is clinically useful, future studies will expand our work to the prediction of different stages of ORNJ to allow for more personalized intervention and management protocols based on the predicted degree of ORNJ severity risk.

Finally, as van den Bosch et al. ^11^ note, there is a significant unmet need for novel higher-dimensional dose-aware toxicity prediction methods to address limitations of standard RT NTCP models as part of an effort to explore non-linear dose-response considerations, and the reality that multiple DVH parameters of the same organ-at-risk (OAR) may be informative have led to applications of “whole DVH” methods ^17^. In this study we used dosimetric variables extracted from DVH data. While DVH is still a widely used surrogate of radiation distribution, it does not incorporate spatial information. As a natural next step from the present work, we aim to combine the proposed time to ORNJ approach to NTCP modeling with spatial information as the dosimetric risk factor in a spatio-temporal ORNJ prediction model.

In conclusion, our ORNJ Weibull AFT (WAFT) model offers a significant advancement in predicting mandibular ORNJ risk following RT in HNC patients. Predicting the time to ORNJ allows for early identification and proactive management of high-risk patients with potential reduction of the severity of ORNJ and improvement of patient outcomes and quality of life.

## Data availability statement

The MDACC deidentified dataset is available in Figshare (10.6084/m9.figshare.26240435). The GSTT dataset was analyzed onsite using a federated method and, due to GDPR restrictions, cannot be made public. Open access/FAIR reporting of scripts and instructions for the model are available in GitHub (https://github.com/LaiaHV-MDACC/ORN-time-to-event-prediction-modelling).

## Acknowledgement

The authors would like to acknowledge the support from various funding sources that made this work possible. C.D.F. and S.Y.L. received related funding support from the NIH/NIDCR (U01DE032168/ R01DE025248). C.D.F. also receives infrastructure and salary support through the NIH/NCI MD Anderson Cancer Center Core Support Grant (CCSG) Image-Driven Biologically-informed Therapy (IDBT) program (P30CA016672-47). S.Y.L. is supported through the CCSG Head and Neck Program (P30CA016672-48). T.G.U. is supported by the Radiation Research Unit at the Cancer Research UK City of London Centre Award [C7893/A28990]. A.C.M. received funding from the NIH/NIDCR via grants K12CA088084, R21DE031082, and K01DE030524. L.V.V. received funding and salary support from KWF Dutch Cancer Society through a Young Investigator Grant (KWF-13529) and from NWO ZonMw through the VENI grant (NWO-09150162010173). K.K.B. acknowledges support from the Image Guided Cancer Therapy Research Program at The University of Texas MD Anderson Cancer Center, which was partially funded by the National Institutes of Health/NCI under award number P30CA016672 and through a generous gift from the Apache Corporation. M.A.N. received funding from the National Institutes of Health/National Institute of Dental and Craniofacial Research (NIH/NIDCR) through grant 1R03DE033550-01. J.R. received salary support from the NIDCR Diversity Supplement Grant 3R01DE028290-02S1. K.A.W. was supported by the Image-Guided Cancer Therapy T32 Training Program Fellowship from T32CA261856. A.J.S. received funding from the NIH/NCI through grant R01CA257814-01. N.A.W. acknowledges funding support from 5R01DE028290-05. G.E.M. acknowledges funding support from NIH UG3 TR004501, NSF CNS-2320261, the University Scholar Award from the University of Illinois System, and NIH/NCI R01CA258827. A.S.R.M. received funding from NIDCR (U01DE032168, 1R01DE028290-01A1) and NCI (R01 CA258827). S.J.F. received funding from Hitachi, NIH/NCI, the National Association of Proton Therapy, Affirmed Pharma and NASA/Baylor College of Medicine and honoraria from IBA.

Additionally, the authors would like to express their gratitude to Brandon Reber, Richard Cardoso and Brandon Gunn for their valuable contributions to this work. Acknowledgements are also made to the following individuals for completing the GUI usability survey: Sigmund Haidacher, Trevor Abts, Rishabh Gaur, Ahmed Tayloun, Sydney Thomas, Sonali Joshi, Irin Luke, Cem Dede.

## References

(1) Frankart, A. J.; Frankart, M. J.; Cervenka, B.; Tang, A. L.; Krishnan, D. G.; Takiar, V. Osteoradionecrosis: Exposing the Evidence Not the Bone. International Journal of Radiation Oncology Biology Physics 2021, 109 (5), 1206–1218. 10.1016/j.ijrobp.2020.12.043.

(2) Laraway, D. C.; Rogers, S. N. A Structured Review of Journal Articles Reporting Outcomes Using the University of Washington Quality of Life Scale. British Journal of Oral and Maxillofacial Surgery 2012, 50 (2), 122–131. 10.1016/j.bjoms.2010.12.005.

(3) O’Dell, K.; Sinha, U. Osteoradionecrosis. Oral Maxillofac Surg Clin North Am 2011, 23 (3), 455–464. 10.1016/j.coms.2011.04.011.

(4) Porcaro, G.; Amosso, E.; Baldoni, M. Treatment of Osteoradionecrosis of the Jaw with Ozone in the Form of Oil-Based Gel: 1-Year Follow-Up. The Journal of Contemporary Dental Practice 2019, 20 (2), 270–276. 10.5005/jp-journals-10024-2508.

(5) Habib, S.; Sassoon, I.; Thompson, I.; Patel, V. Risk Factors Associated with Osteoradionecrosis. Oral Surgery 2021, 14 (3), 227–235. 10.1111/ors.12597.

(6) MD Anderson Head and Neck Cancer Symptom Working Group. Dose-Volume Correlates of Mandibular Osteoradionecrosis in Oropharynx Cancer Patients Receiving Intensity-Modulated Radiotherapy: Results from a Case-Matched Comparison. Radiother Oncol 2017, 124 (2), 232–239. 10.1016/j.radonc.2017.06.026.

(7) Aarup-Kristensen, S.; Hansen, C. R.; Forner, L.; Brink, C.; Eriksen, J. G.; Johansen, J. Osteoradionecrosis of the Mandible after Radiotherapy for Head and Neck Cancer: Risk Factors and Dose-Volume Correlations. Acta Oncol 2019, 58 (10), 1373–1377. 10.1080/0284186X.2019.1643037.

(8) Kubota, H.; Miyawaki, D.; Mukumoto, N.; Ishihara, T.; Matsumura, M.; Hasegawa, T.; Akashi, M.; Kiyota, N.; Shinomiya, H.; Teshima, M.; Nibu, K.-I.; Sasaki, R. Risk Factors for Osteoradionecrosis of the Jaw in Patients with Head and Neck Squamous Cell Carcinoma. Radiat Oncol 2021, 16 (1), 1. 10.1186/s13014-020-01701-5.

(9) Möring, M. M.; Mast, H.; Wolvius, E. B.; Verduijn, G. M.; Petit, S. F.; Sijtsema, N. D.; Jonker, B. P.; Nout, R. A.; Heemsbergen, W. D. Osteoradionecrosis after Postoperative Radiotherapy for Oral Cavity Cancer: A Retrospective Cohort Study. Oral Oncol 2022, 133, 106056. 10.1016/j.oraloncology.2022.106056.

(10) van Dijk, L. V.; Abusaif, A. A.; Rigert, J.; Naser, M. A.; Hutcheson, K. A.; Lai, S. Y.; Fuller, C. D.; Mohamed, A. S. R. Normal Tissue Complication Probability (NTCP) Prediction Model for Osteoradionecrosis of the Mandible in Patients With Head and Neck Cancer After Radiation Therapy: Large-Scale Observational Cohort. Int J Radiat Oncol Biol Phys 2021, 111 (2), 549–558. 10.1016/j.ijrobp.2021.04.042.

(11) Bosch, L. V. den; Schuit, E.; Laan, H. P. van der; Reitsma, J. B.; Moons, K. G. M.; Steenbakkers, R. J. H. M.; Hoebers, F. J. P.; Langendijk, J. A.; Schaaf, A. van der. Key Challenges in Normal Tissue Complication Probability Model Development and Validation: Towards a Comprehensive Strategy. Radiotherapy and Oncology 2020, 148, 151–156. 10.1016/j.radonc.2020.04.012.

(12) Treister, N. S.; Brennan, M. T.; Sollecito, T. P.; Schmidt, B. L.; Patton, L. L.; Mitchell, R.; Haddad, R. I.; Tishler, R. B.; Lin, A.; Shadick, R.; Hodges, J. S.; Lalla, R. V. Exposed Bone in Patients with Head and Neck Cancer Treated with Radiation Therapy: An Analysis of the Observational Study of Dental Outcomes in Head and Neck Cancer Patients (OraRad). Cancer 2022, 128 (3), 487–496. 10.1002/cncr.33948.

(13) Multidisciplinary Larynx Cancer Working Group. Conditional Survival Analysis of Patients With Locally Advanced Laryngeal Cancer: Construction of a Dynamic Risk Model and Clinical Nomogram. Sci Rep 2017, 7, 43928. 10.1038/srep43928.

(14) Wang, S. J.; Lemieux, A.; Kalpathy-Cramer, J.; Ord, C. B.; Walker, G. V.; Fuller, C. D.; Kim, J.-S.; Thomas, C. R. Nomogram for Predicting the Benefit of Adjuvant Chemoradiotherapy for Resected Gallbladder Cancer. J Clin Oncol 2011, 29 (35), 4627–4632. 10.1200/JCO.2010.33.8020.

(15) Wang, S. J.; Kalpathy-Cramer, J.; Kim, J. S.; Fuller, C. D.; Thomas, C. R. Parametric Survival Models for Predicting the Benefit of Adjuvant Chemoradiotherapy in Gallbladder Cancer. AMIA Annu Symp Proc 2010, 2010, 847–851.

(16) Tosado, J.; Zdilar, L.; Elhalawani, H.; Elgohari, B.; Vock, D. M.; Marai, G. E.; Fuller, C.; Mohamed, A. S. R.; Canahuate, G. Clustering of Largely Right-Censored Oropharyngeal Head and Neck Cancer Patients for Discriminative Groupings to Improve Outcome Prediction. Sci Rep 2020, 10 (1), 3811. 10.1038/s41598-020-60140-0.

(17) Hosseinian, S.; Hemmati, M.; Dede, C.; Salzillo, T. C.; van Dijk, L. V.; Mohamed, A. S. R.; Lai, S. Y.; Schaefer, A. J.; Fuller, C. D. Cluster-Based Toxicity Estimation of Osteoradionecrosis Via Unsupervised Machine Learning: Moving Beyond Single Dose-Parameter Normal Tissue Complication Probability by Using Whole Dose-Volume Histograms for Cohort Risk Stratification. Int J Radiat Oncol Biol Phys 2024, 119 (5), 1569–1578. 10.1016/j.ijrobp.2024.02.021.

(18) International ORAL Consortium. International Expert-Based Consensus Definition, Staging Criteria, and Minimum Data Elements for Osteoradionecrosis of the Jaw: An Inter-Disciplinary Modified Delphi Study. medRxiv 2024, 2024.04.07.24305400. 10.1101/2024.04.07.24305400.

(19) Peterson, D. E.; Koyfman, S. A.; Yarom, N.; Lynggaard, C. D.; Ismaila, N.; Forner, L. E.; Fuller, C. D.; Mowery, Y. M.; Murphy, B. A.; Watson, E.; Yang, D. H.; Alajbeg, I.; Bossi, P.; Fritz, M.; Futran, N. D.; Gelblum, D. Y.; King, E.; Ruggiero, S.; Smith, D. K.; Villa, A.; Wu, J. S.; Saunders, D. Prevention and Management of Osteoradionecrosis in Patients With Head and Neck Cancer Treated With Radiation Therapy: ISOO-MASCC-ASCO Guideline. J Clin Oncol 2024, 42 (16), 1975–1996. 10.1200/JCO.23.02750.

(20) Bradburn, M. J.; Clark, T. G.; Love, S. B.; Altman, D. G. Survival Analysis Part II: Multivariate Data Analysis – an Introduction to Concepts and Methods. Br J Cancer 2003, 89 (3), 431–436. 10.1038/sj.bjc.6601119.

(21) Carroll, K. J. On the Use and Utility of the Weibull Model in the Analysis of Survival Data. Controlled Clinical Trials 2003, 24 (6), 682–701. 10.1016/S0197-2456(03)00072-2.

(22) Liu, E.; Liu, R. Y.; Lim, K. Using the Weibull Accelerated Failure Time Regression Model to Predict Time to Health Events. Applied Sciences 2023, 13 (24), 13041. 10.3390/app132413041.

(23) Swindell, W. R. Accelerated Failure Time Models Provide a Useful Statistical Framework for Aging Research. Exp Gerontol 2009, 44 (3), 190–200. 10.1016/j.exger.2008.10.005.

(24) Davidson-Pilon, C. Lifelines: Survival Analysis in Python. Journal of Open Source Software 2019, 4 (40), 1317. 10.21105/joss.01317.

(25) Austin, P. C.; Harrell, F. E.; van Klaveren, D. Graphical Calibration Curves and the Integrated Calibration Index (ICI) for Survival Models. Stat Med 2020, 39 (21), 2714–2742. 10.1002/sim.8570.

(26) Brooke, J. SUS - A Quick and Dirty Usability Scale. In Usability Evaluation In Industry; Taylor & Francis Group: London, 1996.

(27) Orphanet: Osteoradionecrosis of the mandible. https://www.orpha.net/en/disease/detail/521127 (accessed 2024-08-13).

(28) Lang, K.; Held, T.; Meixner, E.; Tonndorf-Martini, E.; Ristow, O.; Moratin, J.; Bougatf, N.; Freudlsperger, C.; Debus, J.; Adeberg, S. Frequency of Osteoradionecrosis of the Lower Jaw after Radiotherapy of Oral Cancer Patients Correlated with Dosimetric Parameters and Other Risk Factors. Head Face Med 2022, 18, 7. 10.1186/s13005-022-00311-8.

(29) Wilde, D. C.; Kansara, S.; Banner, L.; Morlen, R.; Hernandez, D.; Huang, A. T.; Mai, W.; Fuller, C. D.; Lai, S.; Sandulache, V. C. Early Detection of Mandible Osteoradionecrosis Risk in a High Comorbidity Veteran Population. Am J Otolaryngol 2023, 44 (2), 103781. 10.1016/j.amjoto.2022.103781.

(30) Zdilar, L.; Vock, D. M.; Marai, G. E.; Fuller, C. D.; Mohamed, A. S. R.; Elhalawani, H.; Elgohari, B. A.; Tiras, C.; Miller, A.; Canahuate, G. Evaluating the Effect of Right-Censored End Point Transformation for Radiomic Feature Selection of Data From Patients With Oropharyngeal Cancer. JCO Clin Cancer Inform 2018, 2, 1–19. 10.1200/CCI.18.00052.

