## Supplementary Material for "Mandibular dose-volume predicts time-to-osteoradionecrosis in an actuarial normal-tissue complication probability (NTCP) model: External validation of right-censored clinico-dosimetric and competing risk application across international multi-institutional observational cohorts and online graphical"

**Table of Contents**

- **Appendix A.** Transparent Reporting of a Multivariable Prediction Model for Individual Prognosis or Diagnosis (TRIPOD) checklist.
- **Appendix B.** Clinical and dosimetric data for the MD Anderson (internal) and GSTT (external) datasets.
- **Appendix C.** Visual comparison of the actuarial time-to-event (TTE) in months by ORN status for the internal (training and testing) and external datasets.
- **Appendix D.** Multivariable Forward Stepwise Logistic Regression Analysis
- **Appendix E.** Visual analysis of the cumulative hazard function and partial effects of the D25%, dental extraction, and gender variables on survival outcomes.
- **Appendix F.** Results of the external evaluation of the Weibull AFT time-to-event model for ORNJ.
- **Appendix G.** Prospective evaluation of the usability of the ORNJ time-to-event calculator GUI.

**Appendix A. Transparent Reporting of a Multivariable Prediction Model for Individual Prognosis or Diagnosis (TRIPOD) checklist ^A1^.**

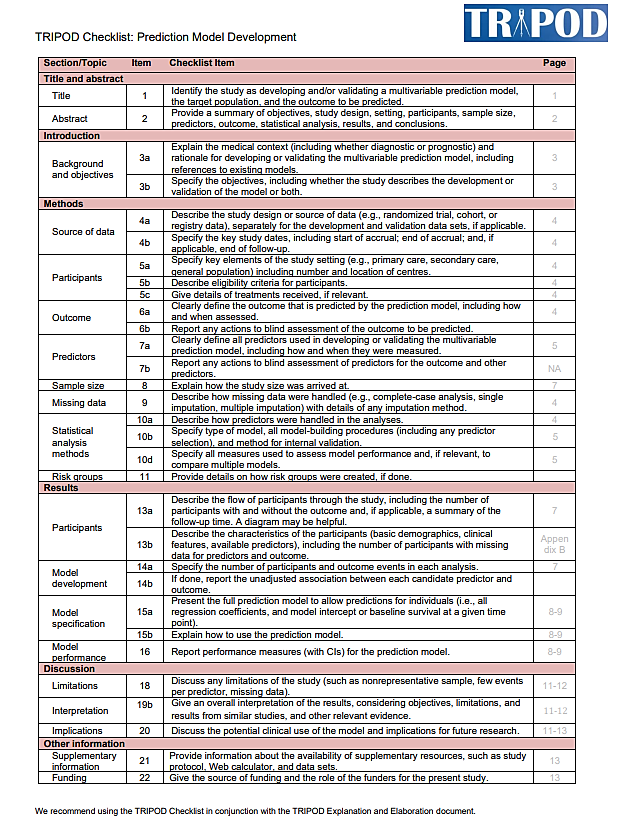

A1. Collins, G. S.; Reitsma, J. B.; Altman, D. G.; Moons, K. Transparent Reporting of a Multivariable Prediction Model for Individual Prognosis or Diagnosis (TRIPOD): The TRIPOD Statement. *BMC Medicine* **2015**, *13* (1), 1. https://doi.org/10.1186/s12916-014-0241-z.

**Appendix B. Clinical and dosimetric data for he MD Anderson (internal) and GSTT (external) datasets.**

*Clinical Data*

Patient clinical and demographic details are summarized in Tables B1 and B2 for the internal and external cohorts, respectively. Of these, smoking status (binary, current vs. never/former), chemotherapy (yes vs. no), gender (binary, male vs. female), pre-RT dental extractions (binary, yes vs. no), post-operative RT (binary, yes vs. no), tobacco pack years (continuous), age (continuous) and mandible volume (continuous) were considered as clinical variables in the multivariable analysis. Only subjects with complete datasets were included. The severity of ORNJ in the training and internal validation cohort was staged using Tsai’s classification ^B1^, as their diagnosis preceded publication of recent consensus severity classification scales ^B2^. ORNJ cases in the external cohort were staged using the Notani staging system ^B3^.

*Dosimetric Data*

The mandibular bone (excluding teeth) was automatically segmented on the planning computed tomography (CT) images using atlas-based approach. The planned radiation dose distribution, mandible structure and CT DICOM files were exported, and an in-house developed Python-based software was used to calculate the dose-volume histogram (DVH) and extract the dose-volume variables. DVH variables included D2%, D5%-D95%, D98% (DX% is the dose received by the most highly irradiated X% of the mandible volume) and V5Gy-V70Gy (VXGy) is the percentage mandible volume receiving at least XGy). For the internal cohort, patients were generally treated with IMRT and prescribed radiation doses ranged between 66 Gy in 30 daily fractions and 70 Gy in 33 daily fractions. For the external cohort, prescribed radiation doses ranged between 50 Gy in 20 daily fractions and 71.5 Gy in 30 daily fractions.

**Table B1. MD Anderson cohort demographic and clinical characteristics**

| **Characteristic** | **Control (N=931)** | **ORN (N=198)** |
| --- | --- | --- |
| **Gender** |  |  |
| Male | 764 (82·1%) | 174 (87·9%) |
| Female | 167 (17·9%) | 24 (12·1%) |
| **Age** (median, IQR) | 60·0 (14·0) | 60·0 (12·0) |
| **Primary tumor site** |  |  |
| Oropharynx | 602 (64·7%) | 138 (69·7%) |
| Oral Cavity | 156 (16·8%) | 49 (24·8%) |
| Larynx/Hypopharynx | 187 (20·1%) | 7 (3·5%) |
| Nasopharynx, Nasal Cavity, Paranasal Sinuses | 21 (2·3%) | 2 (1·0%) |
| Other | 5 (0·5%) | 2 (1·0%) |
| **T stage** |  |  |
| T0 | 33 (3·5%) | 3 (1·5%) |
| T1 | 210 (22·6%) | 31 (15·7%) |
| T2 | 314 (33·7%) | 63 (31·8%) |
| T3 | 200 (21·5%) | 39 (19·7%) |
| T4 | 160 (17·2%) | 59 (29·8%) |
| T4a | 8 (0·9%) | 3 (1·5%) |
| T4b | 0 (0·0%) | 0 (0·0%) |
| TX | 6 (0·6%) | 0 (0·0%) |
| **N stage** |  |  |
| N0 | 189 (20·3%) | 32 (16·2%) |
| N1 | 210 (22·6%) | 30 (15·2%) |
| N2 | 205 (22·0%) | 69 (34·9%) |
| N2a | 23 (2·5%) | 2 (1·0%) |
| N2b | 181 (19·4%) | 40 (20·2%) |
| N2c | 99 (10·6%) | 21 (10·6%) |
| N3 | 21 (2·3%) | 4 (2·0%) |
| NX | 3 (0·3%) | 0 (0·0%) |
| **HPV+** |  |  |
| Yes | 174 (18·7%) | 20 (10·1%) |
| No | 17 (1·8%) | 2 (1·0%) |
| Unknown | 740 (79·5%) | 176 (88·9%) |
| **Smoking status** |  |  |
| Current | 126 (13·5%) | 33 (16·7 %) |
| Former | 445 (47·8%) | 93 (47·0%) |
| Never | 360 (38·7%) | 72 (36·4%) |
| **Pre-RT dental extraction*** | 231 (24·8%) | 76 (38·4%) |
| **Pre-RT surgery** | 66 (9·2%) | 26 (21·0%) |
| Postop RT | 154 (16·5%) | 43 (21·7%) |
| Definitive RT | 777 (83·5%) | 155 (78·3%) |
| **RT technique** |  |  |
| IMRT | 678 (72·8%) | 161 (81·3%) |
| VMAT | 188 (20·2%) | 24 (12·1%) |
| IMPT | 17 (1·8%) | 4 (2·0%) |
| non-IMRT | 11 (1·2%) | 0 (0·0%) |
| NA | 37 (4·0%) | 9 (4·6%) |
| **Chemotherapy** |  |  |
| Concurrent Chemotherapy | 465 (50·0%) | 97 (49·0%) |
| Induction and Concurrent | 202 (21·7%) | 59 (29·8%) |
| No Chemotherapy | 189 (20·3%) | 24 (12·1%) |
| Induction Chemotherapy | 75 (8·1%) | 18 (9·1%) |
| **Follow up time in years** (median, range) | 8·0 (6·6) | 8·0 (5·4) |

*Only dental extractions within six weeks prior to the start date of the radiotherapy course were considered.

**Table B2. GSTT cohort demographic and clinical characteristics**

| Characteristic | Control (N=173) | ORN (N=92) |
| --- | --- | --- |
| **Gender** |  |  |
| Male | 144 (78·3%) | 66 (71·7%) |
| Female | 40 (21·7%) | 26 (28·3%) |
| **Age** (median, IQR) | 61·0 (13·7) | 62·0 (13·0) |
| **Primary tumor site** |  |  |
| Oral cavity | 56 (30·4%) | 28 (30·4%) |
| Oropharynx | 104 (56·5%) | 52 (56·5%) |
| Paranasal sinus/nasopharynx/nasal cavity | 4 (2·2%) | 2 (2·2%) |
| larynx/hypopharynx | 6 (3·3%) | 3 (3·3%) |
| salivary glands | 6 (3·3%) | 3 (3·3%) |
| unknown primary | 8 (4·4%) | 4 (4·4%) |
| **T stage** |  |  |
| T0 | 9 (4·9%) | 4 (4·4%) |
| T1 | 23 (12·5%) | 12 (13·2%) |
| T2 | 62 (33·7%) | 28 (30·8%) |
| T3 | 28 (15·2%) | 9 (9·9%) |
| T4 | 4 (2·2%) | 4 (4·4%) |
| T4a | 56 (30·4%) | 30 (33·0%) |
| T4b | 1 (0·5%) | 4 (4·4%) |
| TX | 1 (0·5%) | 0 (0·0%) |
| **N stage** |  |  |
| N0 | 60 (33·0%) | 25 (27·5%) |
| N1 | 21 (11·5%) | 8 (8·8%) |
| N2a | 15 (8·2%) | 38 (41·8%) |
| N2b | 67 (36·8%) | 38 (41·8%) |
| N2c | 13 (7·1%) | 13 (14·3%) |
| N3 | 5 (2·8%) | 4 (4·3%) |
| NX | 1 (0·6%) | 0 (0·0%) |
| **HPV+** | 56 (83·6%) | 20 (74·1%) |
| **Smoking status** |  |  |
| Current | 50 (27·3%) | 47 (51·1%) |
| Former | 80 (43·5%) | 26 (28·3%) |
| Never | 53 (29·0%) | 19 (20·7%) |
| **Dental extractions** | 113 (65·3%) | 55 (59·8%) |
| **Pre-RT surgery** |  |  |
| Postop RT | 73 (39·7%) | 35 (38·0%) |
| Definitive RT | 111 (60·3%) | 57 (62·0%) |
| **RT technique** |  |  |
| IMRT | 99 (53·8%) | 53 (57·6%) |
| VMAT | 85 (46·2%) | 39 (42·4%) |
| **Chemotherapy** | 110 (59·8%) | 59 (64·1%) |
| **Follow up time in years** (median, range) | 3·9 (2·0) | 4·0 (3·2) |

**Table B3. Tsai and Notani ORNJ classification systems.**

| **Tsai Classification System ^B1^** | |
| --- | --- |
| Stage 1 | Bone exposure or radiographic evidence of bone necrosis without symptoms or infection. |
| Stage 2 | Bone exposure with symptoms such as pain, and possibly secondary infection, but without a pathologic fracture or fistula. |
| Stage 3 | More severe involvement with complications such as a pathological fracture, fistula, or involvement of adjacent structures (e.g., skin or muscles). |
| **Notani Classification System ^B2^** | |
| Stage I | ORN confined to the alveolar bone, which is the part of the jawbone that supports the teeth. |
| Stage II | ORN extending to the mandibular bone, but without pathological fractures. |
| Stage III | ORN with extensive bone involvement, possibly including pathological fractures, or ORN extending beyond the mandibular body (e.g., to the ramus of the mandible or the skull base). |

**References**

B1. Tsai, C. J.; Hofstede, T. M.; Sturgis, E. M.; Garden, A. S.; Lindberg, M. E.; Wei, Q.; Tucker, S. L.; Dong, L. Osteoradionecrosis and Radiation Dose to the Mandible in Patients with Oropharyngeal Cancer. *Int J Radiat Oncol Biol Phys* **2013**, *85* (2), 415–420. https://doi.org/10.1016/j.ijrobp.2012.05.032.

B2. Watson, E. E.; Hueniken, K.; Lee, J.; Huang, S. H.; Maghrabi A, A. E.; Xu, W.; Moreno, A. C.; Tsai, C. J.; Hahn, E.; McPartlin, A. J.; Yao, C. M.; Goldstein, D. P.; De Almeida, J. R.; Waldon, J. N.; Fuller, C. D.; Hope, A. J.; Ruggiero, S. L.; Glogauer, M.; Hosni, A. A. Development and Standardization of a Classification System for Osteoradionecrosis: Implementation of a Risk-Based Model. *medRxiv* **2023**, 2023.09.12.23295454. https://doi.org/10.1101/2023.09.12.23295454.

B3. Notani, K.; Yamazaki, Y.; Kitada, H.; Sakakibara, N.; Fukuda, H.; Omori, K.; Nakamura, M. Management of Mandibular Osteoradionecrosis Corresponding to the Severity of Osteoradionecrosis and the Method of Radiotherapy. *Head & Neck* **2003**, *25* (3), 181–186. https://doi.org/10.1002/hed.10171.

**Appendix C. Visual comparison of the actuarial time-to-event (TTE) in months by ORN status for the internal (training and testing) and external datasets.**

a)

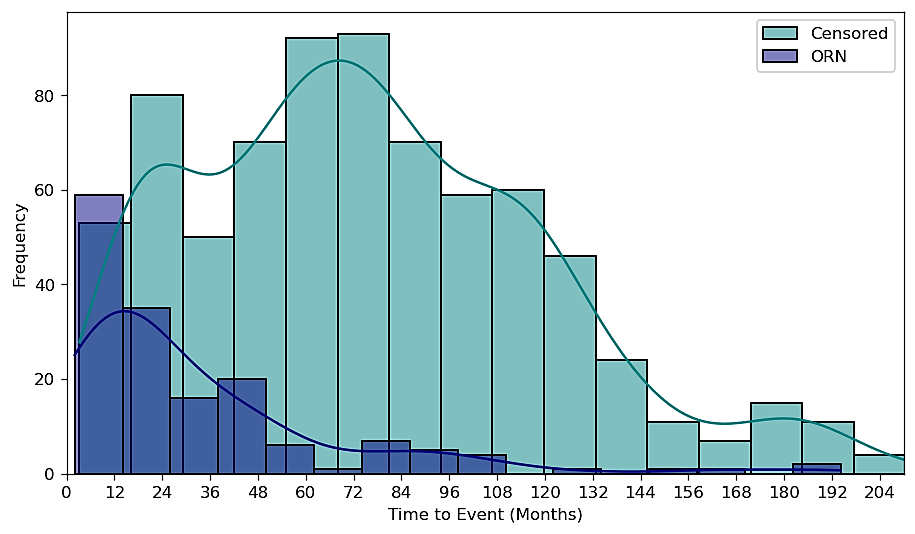

b)

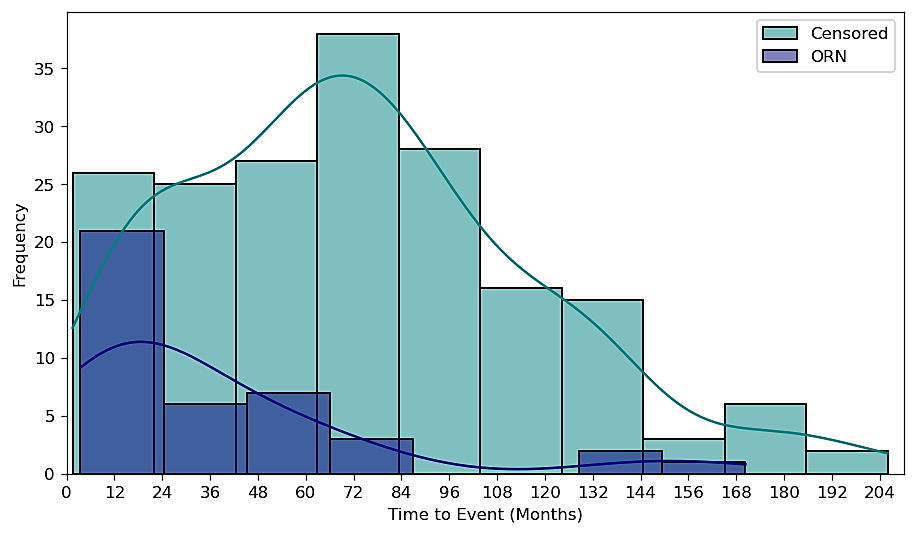

c)

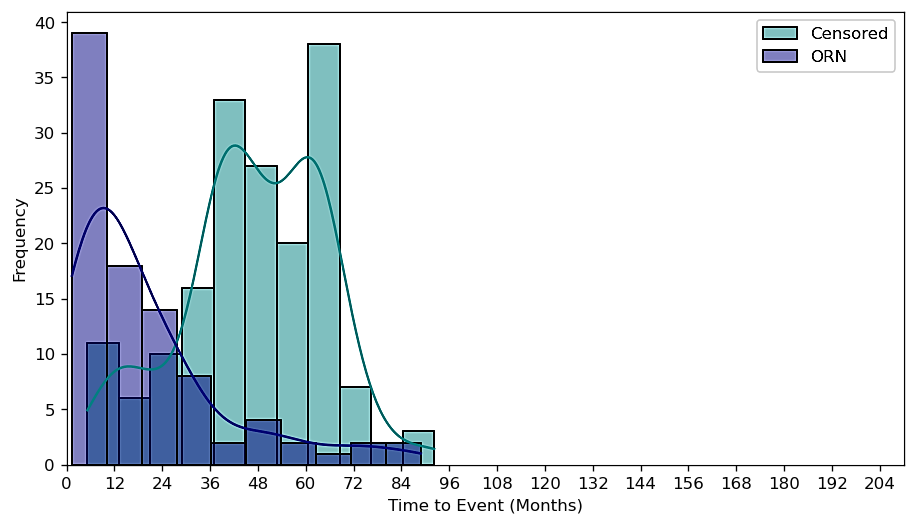

Figure C1. Frequency plot of actuarial time-to-event (TTE) in months by ORN status, where diagnosis of ORN is considered an event censoring either at death or last follow-up, for the training (a) and test (b) internal datasets and for the external dataset (c).

**Appendix D. Multivariable Forward Stepwise Logistic Regression Analysis**

A multivariable logistic regression (LR) analysis with forward stepwise variable selection was carried out in R statistical software using clinical and dosimetric data to identify the most significant variables for prediction of ORNJ to be considered for the Weibull AFT model. The Akaike Information Criterion (AIC) and the Likelihood Ratio Test (LRT) were used as metrics in the forward stepwise variable selection process. The LR model was trained on a training subset of the data (80%) and internally evaluated on the independent test subset (20%), obtained with a random stratified split. A feature pre-selection step was included (on the training subset) based on variable correlation with a Pearson coefficient threshold of 0·8. Bootstrapping (5000 samples) was performed to assess the stability and reliability of the selected variables.

Following the correlation analysis on the initial set of clinical (smoking status, chemotherapy, gender, pre-RT extractions, post-op RT, tobacco pack years, age and mandible volume) and dosimetric variables, the forward stepwise multivariable LR process was first applied considering the pre-selected variables (Figure D1). This identified the following dosimetric and clinical variables as the highest predictors of ORNJ: V55Gy (percentage volume of the mandible that receives at least 55 Gy), D25% (minimum dose received by the most irradiated 25% of the mandible volume), pre-RT dental extractions and gender (supplementary Table D1). However, in this first forward stepwise LR process, the model’s coefficient for the V55Gy parameter was largely altered when the D25% parameter was introduced, showing a correlation between the two dosimetric parameters: V55Gy and D25%. Also, the p-value for the V55Gy showed that this parameter was not statistically significant in the prediction of ORNJ. Consequently, the V55Gy parameter was excluded from the pre-selected variables subset and the modelling process repeated on the updated dataset (supplementary Table D2). Therefore, the final ORNJ NTCP model included D25%, pre-RT dental extractions and gender. Details of the ORNJ NTCP model performance are provided in supplementary Table D3.

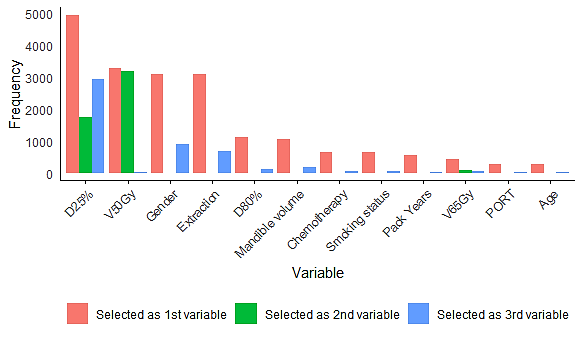

Figure D1. Frequency plot of variable selection during the bootstrapped stepwise forward selection. D25% was the dosimetric variable most frequently selected as first variable, followed by V50Gy. Gender and pre-RT dental extractions were the two clinical variables most frequently selected as first variables.

**Table D1. Models evaluated during the forward stepwise logistic regression process including the V55Gy parameter.** The model coefficient for the V55Gy parameter is largely altered when the D25% parameter is introduced, showing a correlation between the two dosimetric parameters. Also, the p-value for the V55Gy shows that this parameter is not statistically significant in the prediction of ORN. Consequently, the V55Gy was excluded from the variables subset and the modelling process repeated on the updated dataset.

|  | **beta** | **OR** | **p_val** | **AIC** | **BIC** | **p_LRT** |
| --- | --- | --- | --- | --- | --- | --- |
| Intercept | -2·633 | 0·072 |  | 773·9 | 783·5 | < 0·001 |
| V55 | 0·036 | 1·036 | < 0·001 |  |  |  |
| Intercept | -6·745 | 0·001 |  | 747·2 | 761·7 | < 0·001 |
| V55 | 0·001 | 1·001 | 0·864 |  |  |  |
| D25 | 0·094 | 1·099 | < 0·001 |  |  |  |
| Intercept | -5·782 | 0·003 |  | 742·6 | 761·8 | 0·010 |
| V55 | 0·004 | 1·004 | 0·614 |  |  |  |
| D25 | 0·089 | 1·094 | < 0·001 |  |  |  |
| Gender | -0·686 | 0·503 | 0·015 |  |  |  |
| Intercept | -6·296 | 0·002 |  | 739·6 | 763·6 | 0·026 |
| V55 | 0·005 | 1·005 | 0·548 |  |  |  |
| D25 | 0·087 | 1·091 | < 0·001 |  |  |  |
| Gender | -0·655 | 0·520 | 0·020 |  |  |  |
| Dental·Extraction | 0·436 | 1·546 | 0·024 |  |  |  |
| Intercept | -5·985 | 0·003 |  | 739·4 | 768·2 | 0·135 |
| V55 | 0·014 | 1·014 | 0·166 |  |  |  |
| D25 | 0·089 | 1·093 | < 0·001 |  |  |  |
| Gender | -0·668 | 0·513 | 0·018 |  |  |  |
| Dental·Extraction | 0·436 | 1·547 | 0·025 |  |  |  |
| D65 | -0·020 | 0·980 | 0·131 |  |  |  |

**Table D2. Models evaluated during the forward stepwise logistic regression process excluding the V55Gy variable.** The model enclosed in the bold box is the selected ORNJ NTCP model.

|  | **beta** | **OR** | **p_val** | **AIC** | **BIC** | **p_LRT** |
| --- | --- | --- | --- | --- | --- | --- |
| Intercept | -6·861 | 0·001 |  | 745·3 | 754·9 | <0·001 |
| D25% | 0·097 | 1·102 | <0·001 |  |  |  |
| Intercept | -6·144 | 0·002 |  | 740·8 | 755·3 | 0·011 |
| D25% | 0·098 | 1·103 | <0·001 |  |  |  |
| Gender | -0·669 | 0·512 | 0·016 |  |  |  |
| Intercept | -6·723 | 0·001 |  | 738·0 | 757·2 | 0·027 |
| D25% | 0·097 | 1·102 | <0·001 |  |  |  |
| Gender | -0·634 | 0·530 | 0·023 |  |  |  |
| Pre-RT dental extractions | 0·430 | 1·538 | 0·026 |  |  |  |
| Intercept | -7·294 | 0·001 |  | 739·1 | 763·2 | 0·358 |
| D25% | 0·098 | 1·103 | <0·001 |  |  |  |
| Gender | -0·562 | 0·570 | 0·053 |  |  |  |
| Pre-RT dental extractions | 0·421 | 1·523 | 0·030 |  |  |  |
| Mandible volume | 0·005 | 1·005 | 0·358 |  |  |  |

**Table D3. ORNJ NTCP model performance.**

|  | **Training (N=904)** | **Testing (N=225)** |
| --- | --- | --- |
| ROC AUC (95% CI) | 0·760 (0·722-0·760) | 0·698 (0·606-0·698) |
| Nagelkerke R^2^ | 0·195 | 0·115 |
| Brier score | 0·127 | 0·131 |
| Log Loss | 0·403 | 0·418 |
| HL test X^2^ (p-value) | 16·74 (0·03) | 4·52 (0·81) |

**Appendix E. Visual analysis of the cumulative hazard function and partial effects of the D25%, dental extraction, and gender variables on survival outcomes.**

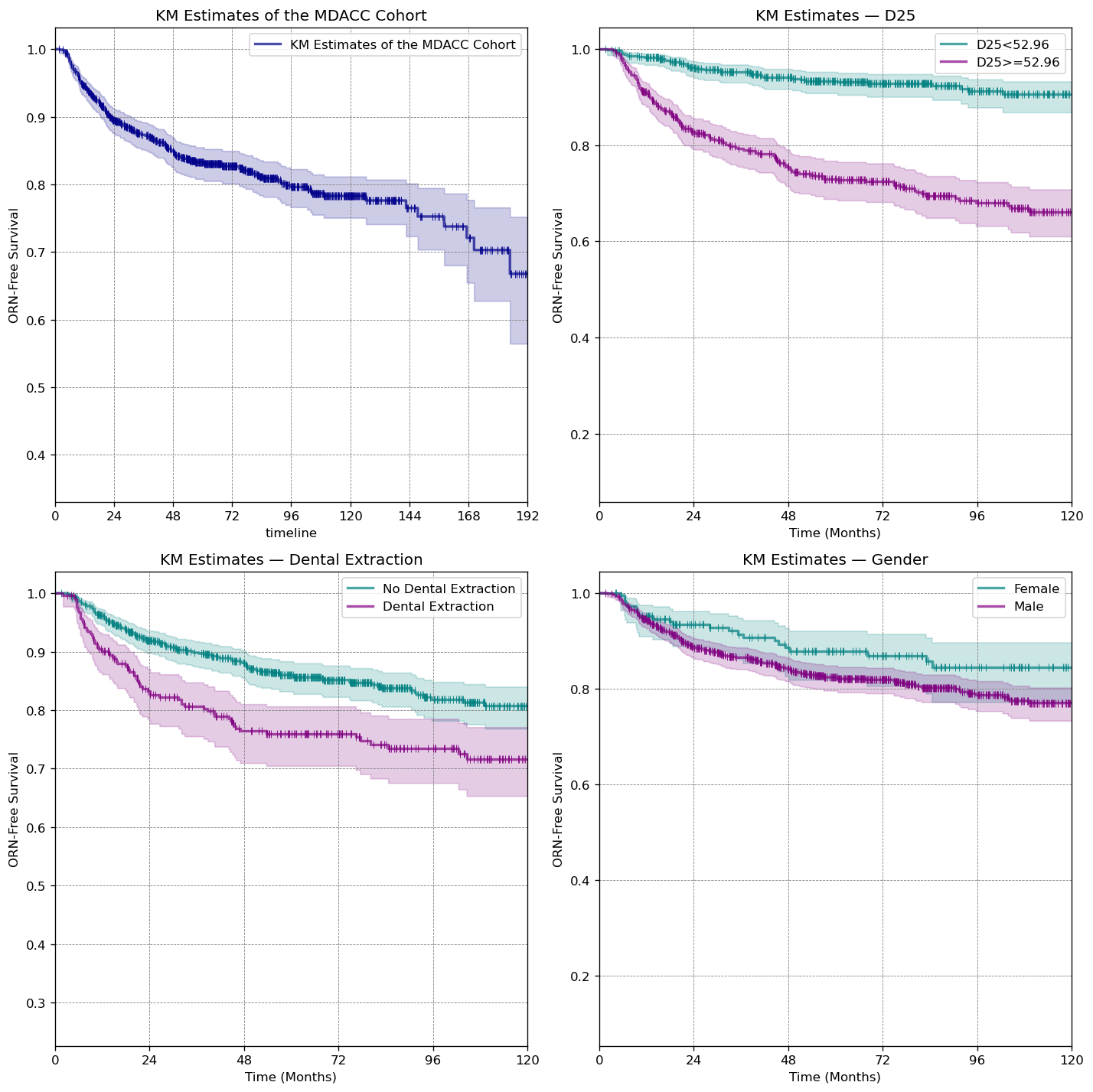

Figure E1. Kaplan-Meier (KM) curves for the entire cohort (top left) and stratified cohorts based on D25% (top right), dental extractions (bottom left) and gender (bottom right).

**Appendix F. Results of the external evaluation of the Weibull AFT time-to-event model for ORNJ.**

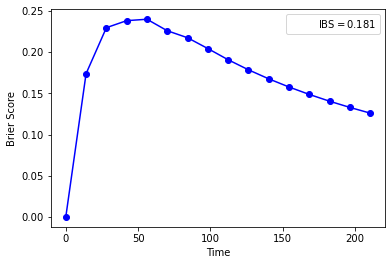

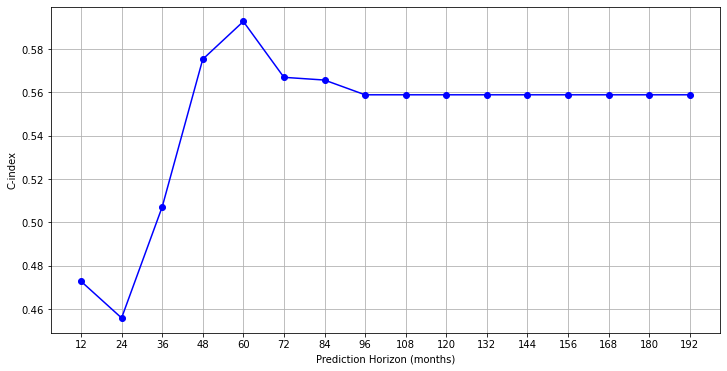

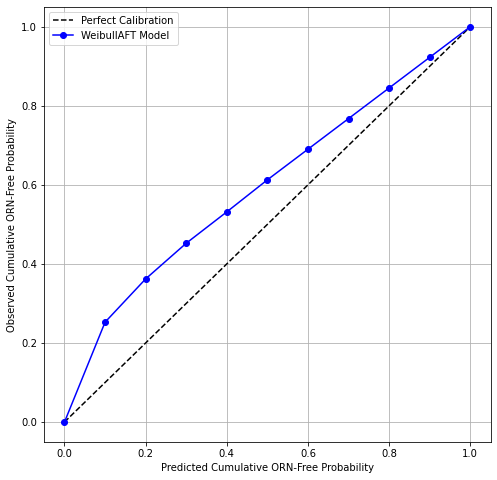

b)

c)

a)

Figure F1: Model performance plots at external validation. Discrimination performance variation over time is described by the Harrell’s C-index (a). Overall model performance over time is described by the Brier score and integrated Brier score (IBS) (b). Model calibration is described by the Distributional calibration curve (c).

**Appendix G. Prospective evaluation of the usability of the ORNJ time-to-event calculator GUI.**

The Qualtrics survey consisted of eight case-specific questions to test the online ORNJ time-to-event calculator GUI, the ten questions from the Brooke et al. (Brooke, 1996) SUS scale questionnaire and 3 additional open questions for additional feedback. The specialties and overall years of clinical experience of the 25 participants that answered the SUS scale questionnaire included 11 students (8 medical, 1 PhD, 1 medical physics, 1 undergrad), Radiation Oncology (5, 0-20 years), Oral Oncology (2, 10-15 years), Oral Surgery (2, 15-20 years), Radiology (1, 5-10 years), Medical Physics (1, 0-5 years), Otolaryngology Head and Neck Surgery (1, 5-10 years), Radiology (1, 5-10 years) and Operations Research (1, 0-5 years). The case-specific questions were answered by 24 out of 25 participants.

**Case-specific questionnaire**

The median overall accuracy in the response of the case-specific questions was 87.5 (84.4), with the lowest accuracy recorded for the Oral Surgery specialty group (Figure G1), which was composed of two specialists with 15-20 years of experience (Figure G2).

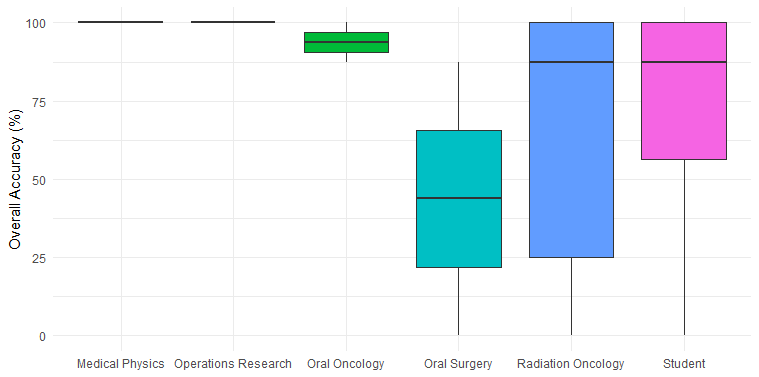

Figure G1. Boxplots of case-specific questionnaire response mean accuracy by specialty group.

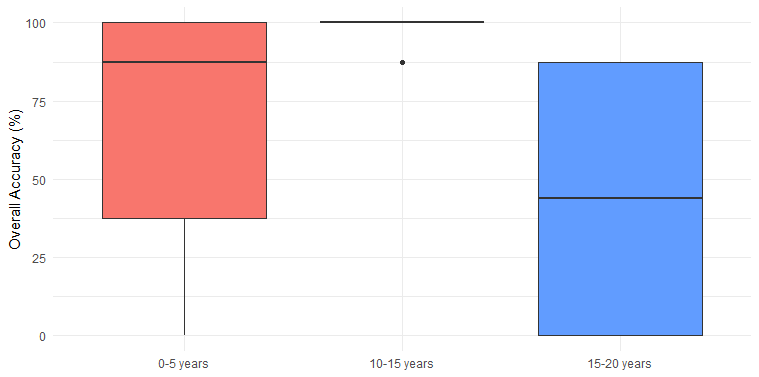

Figure G2. Boxplots of case-specific questionnaire response mean accuracy by years of experience.

Table G1. Percentage of correct responses (accuracy) for case-specific questions across all GUI users.

| **Case-specific questions** | | **Accuracy** |
| --- | --- | --- |
| Case 1: Your patient is a 58 year-old male. His prescribed radiation dose distribution results in a 25% of his mandible (D25%) receiving 57.8 Gy. He had no prior dental extractions prior to initiation of RT. Based on this information: | Q1: What is the predicted risk for ORNJ after 36 months? | 63.3 % |
|  | Q2: What is the predicted risk for ORNJ after 30 months? | 60.0 % |
| Case 2: Your patient is a 66-year old female who had a dental extraction prior to radiation therapy. The patient receives a D25% of 60.69 Gy. Based on this information: | Q3: What is the median time (in months) for expected development of ORNJ for prior patients with these parameters? | 56.7 % |
|  | Q3: If this patient’s gender were Male, would the patient be expected to be at greater risk to develop ORNJ earlier or later than the equivalent female patient? | 66.7 % |
| Case 3: Your patient is a 57-year old male who has not received a prior dental extraction. The patient receives a D25% of 39.59 Gy. Based on this information: | Q5: What is the patient’s population-based predicted risk of ORNJ at 24 months? | 66.7 % |
|  | Q6: Assuming the same dose (D25%=39.59 Gy) applies, what is this patient-equivalent population-based expected mean ORNJ-free time in months? | 53.3 % |
| Case 4: Your patient is a 47-year old female with a history of previous dental extraction. The patient receives a D25% of 63.74 Gy. Based on this information: | Q7: What is the patient’s population-based predicted risk of ORNJ after 55 months? | 53.3 % |
|  | Q8: For this same patient, did dental extractions increase the risk of progression to ORNJ at the stated time-point (55 months after RT)? | 66.7 % |

**System Usability Scale (SUS) score**

The System Usability Scale (SUS), developed by John Brooke in 1986 ^26^, is a widely accepted standard for assessing the usability of systems and products. Comprising 10 questions, SUS captures users' experiences by having them rate each item on a five-point Likert scale, ranging from strong disagreement (1) to strong agreement (5). The questions are designed to alternate between positive and negative statements to minimize bias and ensure balanced feedback. The SUS score can range from 0 to 100, where higher scores indicate better usability (a score above 69 is considered above average usability). Each item on the survey (Q1-Q10) is rated individually and the corresponding SUS scores are calculated as follows:

For the odd-numbered questions (Q1, Q3, Q5, Q7, Q9): Score_SUS_ = Rating – 1. For the even-numbered questions (Q2, Q4, Q6, Q8, Q10): Score_SUS_ = 5 – Rating. The final SUS score is obtained by adding all 10 individual scores and multiplying it by 2.5 to convert it to a scale of 0 to 100:

$$SUS score=2.5 \times\sum_{i=1}^{10} \left( {Score}_{SUS} for Q_{i} \right)$$

The overall average SUS score received was 85.0 (40.0-100.0); the median score was 82.5 (IQR 28.8). Except for the Oral Surgery group (mean SUS score 57.5, range 55.0-60.0) all other specialty groups scored the usability GUI above average (Figure G3). The SUS score increased with number of years of experience (Figure G4) except for the group with 15-20 years of experience; this group was composed of four participants, two of which were the Oral Surgeons in the survey. Interestingly, the Oral Surgery specialty group scored the lowest case-specific response accuracy across all specialty groups (Figure G1). Unfortunately, no specific feedback was received from these two users. Low SUS scores were also obtained from two other participants, a medical student (40.0) who reported limited background knowledge as a limitation in the use of the GUI and a Radiation Oncologist with 0-5 years of experience (52.5).

Table G2. Question-specific rating and adjusted score received for the Brooke et al. ^26^ SUS scale questionnaire.

| **SUS question ^26^** | **Median rating (IQR)** | **Median score (IQR)** |
| --- | --- | --- |
| Q1: Would use system frequently | 4 (1) | 3 (1) |
| Q2: Found system unnecessarily complex | 2 (2) | 3 (2) |
| Q3: System was easy to use | 4 (1) | 3 (1) |
| Q4: Would need technical support to use system | 2 (2) | 3 (2) |
| Q5: System functions were well integrated | 4 (1) | 3 (1) |
| Q6: Too much inconsistency in the system | 2 (1) | 3 (1) |
| Q7: Most people would learn to use the system quickly | 4 (1) | 3 (1) |
| Q8: System was cumbersome to use | 1 (1) | 4 (1) |
| Q9: Felt very confident to use the system | 4 (1) | 3 (1) |
| Q10: Much to learn before using the system | 2 (2) | 3 (2) |

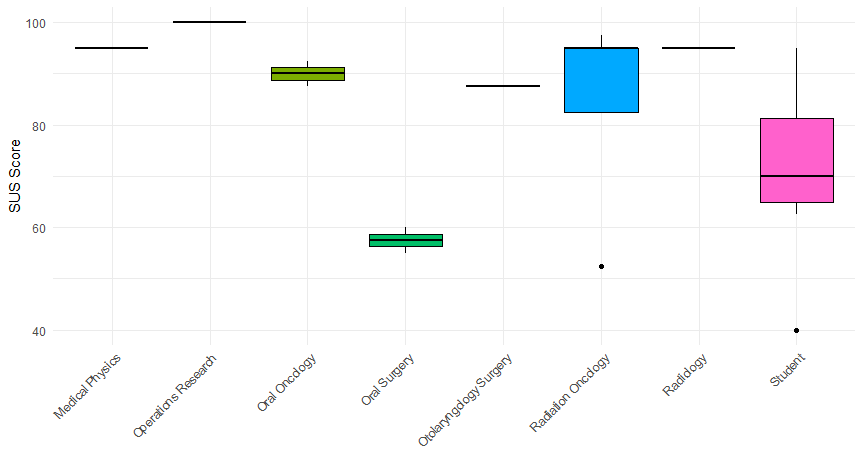

Figure G3. Boxplots of the SUS scores obtained by specialty groups. A score above 69 is considered above average usability.

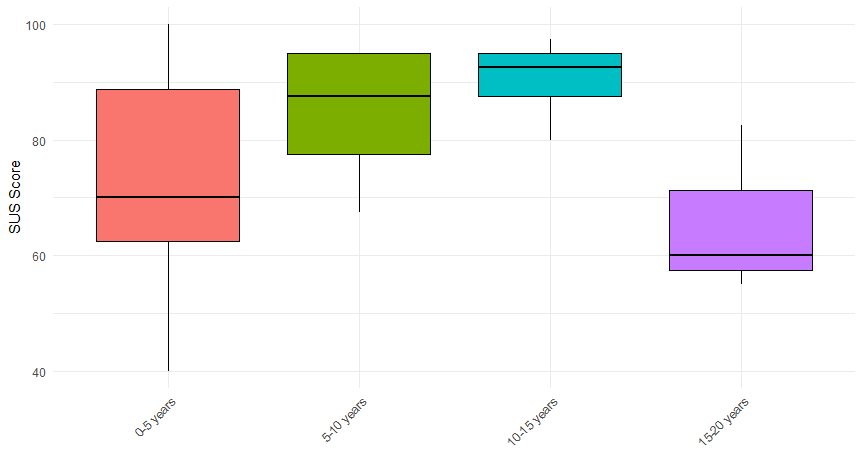

Figure G4. Boxplots of the SUS scores obtained by years of experience. A score above 69 is considered above average usability.

**Additional feedback on GUI aesthetics and usability**

The feedback on the system’s aesthetics and design revealed both strengths and areas for improvement. Users appreciated the overall organization and visual appeal of the system, including its curves, lines, and font choices. However, many found the terminology and components confusing due to limited background knowledge. To address this, it was suggested to include detailed instructions and a README to clarify each part of the interface, along with interactive explanations or tooltips. Additionally, the colors of the scroll bars were noted as less favorable, and it was recommended to update them to match the design better and to use curved boxes for a modern look. Functional enhancements such as adding a "Print" button, an "Email Report" feature, and a quick feedback system were also proposed to improve usability and convenience. Overall, while the system was recognized for its simplicity and efficiency, incorporating these changes would enhance user understanding and functionality.

Overall, the tool was appreciated for its ease of use, but, according to the survey participants, improvements in terminology, layout, response validation, and customization options would enhance its functionality and user experience. For instance, there was confusion around the term "survival," especially regarding patients before and after developing Osteoradionecrosis (ORN). The wording might need adjustment for better understanding. Another suggestion received was that adding an option to change displayed times in the top right box would enhance usability, allowing for adjustments based on clinical needs.
